# Associated socio-demographic and psychological factors of childhood overprotection/overcontrol experiences among Chinese university students: a nationwide survey

**DOI:** 10.1101/2023.04.13.23288506

**Authors:** Jiamei Zhang, Zhipeng Wu, Haojuan Tao, Min Chen, Miaoyu Yu, Liang Zhou, Meng Sun, Dongsheng Lv, Guangcheng Cui, Qizhong Yi, Hong Tang, Cuixia An, Zhening Liu, Xiaojun Huang, Yicheng Long

## Abstract

The childhood experiences of being overprotected and overcontrolled by family members have been suggested to be potentially traumatic. However, the possible associated factors of these experiences among young people are still not well studied. This study aimed to investigate the possible associated factors of childhood overprotection/overcontrol (OP/OC) experiences in young populations in a relatively large, nationwide sample of Chinese university students. A total of 5,823 university students across nine different provinces in China were recruited and included in the data analyses. All participants completed the OP/OC subscale in a recently developed 33-item Childhood Trauma Questionnaire (CTQ-33) to assess their OP/OC experiences. Data were also collected on socio-demographic information and multiple psychological characteristics of all participants. Binary logistic regression was conducted to investigate the associated factors of OP/OC. The prevalence of childhood OP/OC was estimated as 15.63% (910/5,823) based on a cutoff of OP/OC subscale score ≥ 13. Binary logistic regression suggested that being male, being a single child, having depression, having psychotic-like experiences, lower family functioning, and lower psychological resilience were independently associated with childhood OP/OC experiences (all corrected-*p*<0.05). The OP/OC was also positively associated with all the other trauma subtypes (abuses and neglects) in the CTQ-33. Post-hoc analyses suggested that OP/OC experiences were associated with depression in only females, and associated with anxiety in only males. Our results may provide initial evidence that childhood OP/OC experiences would have negative effects on young people’s mental health which merits further investigations, especially in clinical populations.

## 1. Introduction

Overprotection/overcontrol (OP/OC) behaviors were defined as behaviors in which caregivers (including parents and other family members) are overly involved in children’s daily activities and experiences, often caused by excessive anxiety about the children’s safety (Şar et al., 2021; Wood et al., 2003). There are multiple possible reasons which may lead to OP/OC behaviors: for example, some parents exhibited fear in fulfilling their parenting responsibilities, which may in turn leading to their OP/OC (Holmbeck et al., 2002); that lack of care by one parent can also lead to OP/OC by the other (Cella et al., 2014). Prior research has shown that perceived OP/OC from family members might limit children to develop a clear understanding of environmental dangers and might have negative effects on their mental health statuses (Affrunti & Woodruff-Borden, 2015; Miller et al., 2018). For instance, perceived OP/OC experiences were suggested to be possibly associated with decreased self-efficacy and increased vulnerability to perceived threats (Wood et al., 2003), the development of childhood anxiety (Holmbeck et al., 2002), as well as the onset of anorexia (Albinhac et al., 2019) in children and teenagers. In addition, OP/OC might be related to increased risks of depression, post-traumatic stress disorder (Williamson et al., 2017), and suicidal behaviors (Goschin et al., 2013).

In addition to the short-term negative psychological effects of OP/OC in children/teenagers as mentioned above, recent studies have also suggested that OP/OC might be developmentally traumatizing, and childhood OP/OC experiences may have long-term effects on one’s mental health in early adulthood and even later life (Affrunti & Woodruff-Borden, 2015; Lima et al., 2010; Vigdal & Brønnick, 2022). For instance, individual recall of childhood OP/OC appears to be associated with higher prevalence and incidence of adult psychological health problems in the general population (McLafferty et al., 2019; Overbeek et al., 2007; Şar & Türk-Kurtça, 2021). Some evidence suggested that childhood OP/OC experiences are related to sleep disturbance (Shibata et al., 2016) and associated with difficult recovery in patients with schizophrenia (Ishii et al., 2020) in adulthood. Another related study reported that overprotective support reduced stress in the short term, but hinder individuals from coping with stress in the long run by weakening autonomy, especially when that support is terminated (Zniva et al., 2017). For these reasons, perceived OP/OC during childhood has been regarded as a kind of traumatic experiences besides the other well-known childhood trauma subtypes (e.g., abuses and neglects) and attracted attentions in recent psychological studies (Şar et al., 2021; Wu, Liu, Jiang, et al., 2022). Recognizing and identifying factors associated with childhood OP/OC experiences, therefore, may be valuable for improving our understanding of the developments of common mental problems and disorders, as well as finding potential targets for early interventions for mental disorders.

The current literatures about possible associated factors of childhood OP/OC experiences in young people, however, are still limited in several ways. Firstly, some previous results have reported inconsistent and even conflicting conclusions. For example, while many earlier studies as mentioned above suggested that childhood OP/OC is related to more mental problems including depressive and anxiety symptoms in later life, the opposite results were also reported, e.g., that paternal overcontrols predicted lower anxious-depressed symptoms (Basili et al., 2021). One of the potential reasons for these contradictory results may be the insufficient sample size in many of these studies; for instance, the samples in most of the previous studies range from only dozens to hundreds (Bark et al., 2021; Hemm et al., 2018; Spada et al., 2012; Van Petegem et al., 2022), which may lead to a relatively low statistical power and unreliable results. Secondly, most of the prior studies have focused on the associations between OP/OC and several common mental problems such as anxiety and depression; however, the knowledge is limited on the relationships between OP/OC and some other important socio-demographic and psychological characteristics. These characteristics include, for example, psychological resilience which is defined as one’s ability to recover and maintain adaptive behaviors when facing constant stress (Long et al., 2019). There has been evidence that other subtypes of childhood trauma (e.g., abuses and neglects) could lead to a lower psychological resilience, which mediates the relationships between childhood trauma and depression in college students (Chang et al., 2021). As a kind of traumatic experiences, OP/OC experiences may be also associated with a lower psychological resilience, which remains however poorly investigated to our knowledge. Thirdly, while most of the prior studies on OP/OC experiences were conducted in western countries, it is relatively little known about the prevalence and associated factors of OP/OC among youths under other culture conditions, such as in China. One possible reason for such a limitation is the lack of an easy and feasible screening tool for OP/OC experiences in Chinese language before. Nevertheless, this gap has been addressed by a recently validated Chinese version of the 33-item expanded Childhood Trauma Questionnaire (CTQ-33) (Wu, Liu, Jiang, et al., 2022), and further studies on OP/OC among young Chinese populations may be warranted.

In the current study, we aim to address the limitations raised above by performing a nationwide, large-sample survey among young Chinese population. In specific, data on childhood OP/OC experiences and multiple socio-demographic/psychological factors (e.g., psychological resilience) were collected from a total of 5,823 Chinese university students across nine different provinces in China. Logistic regression models were conducted to investigate the possible associations between childhood OP/OC experiences and other socio-demographic/psychological factors. We hope that our results would shed light on the understanding of possible role of OP/OC in psychological health among young people.

## 2. Methods

### 2.1. Participants

A total of 5,993 Chinese university students were initially recruited in this survey from nine universities across nine different provinces (Shandong, Jiangxi, Guangxi, Guangdong, Hebei, Inner Mongolia, Heilongjiang, Hunan, and Xinjiang) in China (see distributions in **Table 1**). The survey was conducted during September 2021-October 2021, and all students completed the survey online through a famous platform in China, “Questionnaire Star” (www.wjx.xn). To avoid the potential confounding impacts of other clinical conditions on the results, students with a previous diagnosis of any psychiatric disorder were excluded (*n* = 120). In addition, students with missing data (*n* = 47) or over the age of 25 (*n* = 3) was excluded. Therefore, 5,823 participants were included in the final data analyses in the current study (see **Table 1** for sample characteristics). All participants and/or their guardians gave informed consent to agree to participate in this study. The research proposal was approved by the Ethics Committee of the Second Xiangya Hospital of Central South University.

**Table 1.** Sample characteristics of the analyzed participants.

| University | Located province | Number of participants | Females (%) / males | Age (mean ± SD) | Scores of CTQ-OP/OC subscale | | |
| --- | --- | --- | --- | --- | --- | --- | --- |
|  |  |  |  |  | Mean | SD | Mean + SD |
| Jining Medical University | Shandong | 1212 | 694(57.26)/518 | 19.56 ± 1.28 | 8.88 | 3.64 | 12.52 |
| Gannan Medical university | Jiangxi | 3065 | 1717(56.02)/1348 | 19.50 ± 1.56 | 9.01 | 3.37 | 12.38 |
| Guangxi Medical university | Guangxi | 317 | 187(58.99)/130 | 19.96 ± 1.31 | 8.99 | 3.18 | 12.17 |
| Guangzhou Medical university | Guangdong | 85 | 51(60.00)/34 | 19.49 ± 1.36 | 9.28 | 3.14 | 12.42 |
| Hebei Medical university | Hebei | 161 | 105(65.22)/56 | 18.25 ± 0.74 | 8.61 | 3.08 | 11.69 |
| Inner Mongolia Medical University | Inner Mongolia | 131 | 100(76.34)/31 | 18.73 ± 0.92 | 8.15 | 3.41 | 11.56 |
| Qiqihar Medical University | Heilongjiang | 298 | 177(59.40)/121 | 18.37 ± 0.79 | 8.62 | 3.49 | 12.11 |
| Central South University | Hunan | 394 | 210(53.30)/184 | 18.28 ± 1.08 | 9.08 | 3.48 | 12.56 |
| Xinjiang Medical university | Xinjiang | 160 | 102(63.75)/58 | 20.01 ± 1.41 | 8.56 | 3.57 | 12.13 |
| Total | | 5823 | 3343(57.41)/2480 | 19.36 ± 1.47 | 8.93 | 3.43 | 12.36 |

Note: CTQ, Childhood Trauma Questionnaire; OP/OC, overprotection/overcontrol; SD, standard deviation.

### 2.2. Assessments

#### 2.2.1. Socio-demographic information

Information on the following socio-demographic factors were collected from all participants and taken into the analyses: age, sex, ethnicity (Han or minority), single-child household (yes or no), parental separation (yes or no), left-behind children experiences (yes or no), as well as family histories of mental disorders. Note that all participants with personal history of mental disorders have been excluded from the analyses.

#### 2.2.2. Measure of OP/OC experiences

Childhood OP/OC experiences of all participants were measured by the OP/OC subscale of the CTQ-33 (Şar et al., 2021). The CTQ-33 was expanded from the original 28-item Childhood Trauma Questionnaire (CTQ-28) (Bernstein et al., 1994) with an additional OP/OC subscale, and thus has six subscales measuring six different subtypes of childhood trauma experiences: emotional abuse, physical abuse, sexual abuse, physical neglect, emotional neglect, and OP/OC (Şar et al., 2021). All items in the CTQ-33 are 5-point Likert-type questions, and higher scores indicate higher levels of childhood trauma experiences. The Chinese version of the original CTQ-28 has been shown to have good reliability and validity in Chinese populations (Xiang et al., 2021). The additional OP/OC subscale in the CTQ-33 has also been translated into Chinese and proved to be valid (Wu, Liu, Jiang, et al., 2022).

Based on prior publications, participants with scores above the cutoff points for a particular subscale can be defined as having a particular subtype of childhood trauma experience as follows: physical abuse ≥ 10, emotional abuse ≥ 13, sexual abuse ≥ 8, physical neglect ≥ 10, and emotional neglect ≥ 15 (D. Huang et al., 2021; Tietjen et al., 2010). In the present study, we intended to firstly classify all participants into those with and without childhood OP/OC experiences. However, to our knowledge, an optimal cutoff point for the OP/OC subscale in the CTQ-33 has not been established to date. Therefore, referring to multiple published studies (Asarnow et al., 2019; Liu et al., 2022; Quintero Garzón et al., 2021), we estimated the appropriate cutoff score for the OP/OC subscale based on one standard deviation (SD) above the mean score in the surveyed sample. The participants with a OP/OC subscale score higher than such a cutoff point were then defined as having childhood OP/OC experiences.

#### 2.2.3. Self-reported depression

All participants completed the self-reported, 9-item Patient Health Questionnaire (PHQ-9) (Kroenke et al., 2001) to assess the severity of depressive symptoms over the past two weeks. The Chinese version of PHQ-9 has been validated in previous study (Wang et al., 2014). Each item of the PHQ-9 was rated on 4 values ranging from 0 (“not at all”) to 3 (“nearly every day”). The total score of PHQ-9 can range from 0 to 27, and the participants were regarded to have depression when the total score >=10 (Levis et al., 2019).

#### 2.2.4. Self-reported anxiety

All participants completed the self-reported, 7-item Generalized Anxiety Disorder Scale (GAD-7) (Spitzer et al., 2006) to assess their anxiety levels during the last two weeks. The Chinese version of GAD-7 has shown good reliability and validity in Chinese populations (Tong et al., 2016; Zhang et al., 2021). Each item of the GAD-7 was rated from 0 (“not at all”) to 3 (“nearly every day”). The total score of GAD-7 can range from 0 to 21, and the participants were regarded to have anxiety when the total score >=10 (Tong et al., 2016; Zhang et al., 2021).

#### 2.2.5. Psychotic-like experiences

The 15-item version of the Community Assessment of Psychic Experiences (CAPE-15) (Konings et al., 2006; Sun et al., 2020; Zhang et al., 2022) was used to evaluate the psychotic-like experiences for all participants. The Chinese version of the CAPE has been validated and is widely used to assess psychotic-like experience in Chinese populations (Sun et al., 2017; Wang et al., 2022; Wu, Jiang, et al., 2022; Wu, Liu, et al., 2021). The CAPE-15 include 15 items which measured both frequency of and distress associated with a series of common psychotic-like experiences (e.g., subclinical delusions and hallucinations). Both the frequency and distress scores of each item are rated on a four-point Likert scale. Referring to prior studies (Bukenaite et al., 2017), the participants were regarded to have meaningful psychotic-like experiences when both the mean frequency score and mean distress score are greater than 1.5.

#### 2.2.6. Family functioning

The family functioning of each participant was measured by the Family APGAR scale (Smilkstein, 1978; Wu, Zou, et al., 2021). The Chinese version of Family APGAR has been validated and widely used in previous studies (Hai et al., 2017; Y. Huang et al., 2021; Wu, Zou, et al., 2021). The Family APGAR scale consists of 5 items assessing family functioning from five dimensions: adaptation (“A”), partnership (“P”), growth (“G”), affection (“A”), and resolution (“R”). The score of each item ranges from 0 (“almost always”) to 2 (“hardly ever”). Total scores of the Family APGAR scale can thus range from 0 to 10, and a relatively low family functioning can be defined by a total score <=3 (Hai et al., 2017; Y. Huang et al., 2021; Wu, Zou, et al., 2021).

#### 2.2.7. Psychological resilience

Each participant’s psychological resilience was measured by the 10-item Connor-Davidson Resilience Scale (CDRS-10) (Campbell-Sills & Stein, 2007), a self-administered questionnaire extracted from the original 25-item CDRS (Connor & Davidson, 2003). The Chinese version of CDRS has been validated and widely used in previous studies (Guo et al., 2021; Long et al., 2019; Ye et al., 2017). In the CDRS-10, the score of each item ranges from 0 to 4 (0 = “never” to 4 = “almost always”), and the total score ranges from 0 to 40. Referring to prior research (Ye et al., 2017), the cutoff of a CDRS-10 total score <=25 was used to define a relatively low psychological resilience.

### 2.3. Statistical analyses

Socio-demographic and psychological characteristics were firstly compared between the participants with and without childhood OP/OC experiences using descriptive statistics. Independent *t*-tests and chi-square tests were used for continuous variables (e.g., age) and categorical variables (e.g., sex), respectively.

In line with some prior studies (Wu, Liu, Zhang, et al., 2022), binary logistic regression analysis was then performed to investigate the possible associations between all socio-demographic/psychological factors (age, sex, years of education, ethnicity, province, single child, parental separation, left-behind experiences, family history of mental disorders, depression, anxiety, psychotic-like experiences, family functioning, and psychological resilience) and childhood OP/OC experiences after adjusting for the confounding effects of other variables. The statistical significance was set at *p* < 0.05 (two-tailed) after false discovery rate (FDR) corrections. Note that the province (coded as dummy variables) was controlled in the analyzing model as a variable of no interest. Moreover, since previous studies have suggested that OP/OC is highly positively correlated with all the other trauma subtypes (abuses and neglects) in the CTQ-33 (Şar et al., 2021), we didn’t include other subscales of the CTQ-33 in the regression model to avoid possible multicollinearity problems. Instead, we explored their relationships with OP/OC using separate models in the following supplementary analyses (see later in **Section 2.4**).

### 2.4. Supplementary analyses

Several supplementary analyses were performed in addition to the main analyses. Firstly, we tested the relationships between OP/OC and other trauma subtypes (abuses and neglects) in the CTQ-33 using Spearman correlations. We also tested whether OP/OC and other trauma subtypes would have similar associated socio-demographic/psychological factors: here, all participants were classified into those with and without a particular subtype of childhood trauma (e.g., psychical abuse, based the cutoffs mentioned in **Section 2.2.2**), and separate binary regression models were used to investigate the associated factors of such trauma subtype. Similar to the analyses on OP/OC, the statistical significance was set at FDR-corrected *p* < 0.05.

Secondly, considering that sex differences in mental health have been widely reported (Maeng & Milad, 2015; Rubinow & Schmidt, 2019; Wu, Wang, et al., 2022), we further explored the possible sex differences in relationships between OP/OC and other factors. Here, similar to analyses in the entire sample, the associated factors of childhood OP/OC experiences were assessed by binary logistic regression models in the male (*N* = 2,480) and female (*N* = 3,343) participants separately, and the statistical significance was still set at FDR-corrected *p* < 0.05.

### 2.5. Validation analysis

In the current study, we estimated an appropriate cutoff point for the OP/OC subscale at >= 13. To confirm whether the identified associated factors of OP/OC would change when using different cutoff scores, we repeated the regression analyses using two other different cutoff points (>= 12 and >= 14, respectively).

## 3. Results

### 3.1. Sample characteristics and estimated cutoff points

Sample characteristics of the analyzed participants are shown in **Table 1**. The ratio of female participants was 57.41% (3,343/5,823), and the average age was 19.36 (SD = 1.47) for the entire sample. The (mean + SD) values of the OP/OC subscale scores were found to be very close across the subsamples from different provinces: in most (7/9) of the subsamples, such values were in the range of 12.11-12.56; only two exceptions were found in Hebei (11.69) and Inner Mongolia (11.56), two provinces with relatively small sample size (*N* = 161/131 for Hebei and Inner Mongolia, respectively). Meanwhile, the (mean + SD) values of the OP/OC subscale score was 12.36 when calculated in the entire sample. According to such results, we thus propose that an OP/OC subscale score of ≥ 13 may be an appropriate cutoff to classify those having and not having clinically meaningful OP/OC experiences. Such a cutoff point was also applied in the following analyses.

### 3.2. Group comparisons on socio-demographic and psychological characteristics

Based on the above cutoff point (OP/OC subscale score ≥ 13), the prevalence of OP/OC experiences was estimated as 15.63% (910/5,823) in the current sample. Results of the direct comparisons on all socio-demographic and psychological characteristics between the participants with and without OP/OC experiences were shown in **Table 2**. Compared with those without OP/OC, the participants with OP/OC experiences had a higher proportion of males (*p* < 0.001), a higher proportion of single child (*p* = 0.031), and a higher proportion of “left-behind” children (*p* = 0.006). Compared with those without OP/OC, the participants with OP/OC experiences are more likely to have depression, anxiety, psychotic-like experiences, low family functioning, and low psychological resilience (all *p* < 0.001).

**Table 2.** Comparisons on socio-demographic and psychological characteristics between the participants with and without childhood OP/OC experiences.

| Variables | With OP/OC (*n* = 910) | Without OP/OC (*n* = 4913) | Group comparison |
| --- | --- | --- | --- |
| Age (years, mean ± SD) | 19.31 ± 1.48 | 19.37 ± 1.47 | $t = 1.064, p = 0.287$ |
| Sex | | | $\chi^2 = 115.786, p < 0.001^{***}$ |
| Male, *n* (%) | 535 (58.79%) | 1945 (39.59%) |  |
| Female, *n* (%) | 375 (41.21%) | 2968 (60.41%) |  |
| Years of education (mean ± SD) | 13.09 ± 1.25 | 13.11 ± 1.19 | $t = -0.482, p = 0.630$ |
| Ethnicity | | | $\chi^2 = 0.079, p = 0.779$ |
| Han, *n* (%) | 851 (93.52%) | 4582 (93.26%) |  |
| Minority, *n* (%) | 59 (6.48%) | 331 (6.74%) |  |
| Single child, *n* (%) | 300 (32.97%) | 1444 (29.40%) | $\chi^2 = 4.679, p = 0.031^*$ |
| Parental separation, *n* (%) | 98 (10.77%) | 500 (10.18%) | $\chi^2 = 0.292, p = 0.589$ |
| Left-behind experiences, *n* (%) | 318 (34.95%) | 1492 (30.37%) | $\chi^2 = 7.507, p = 0.006^{**}$ |
| FHMD, *n* (%) | 17 (1.87%) | 78 (1.59%) | $\chi^2 = 0.376, p = 0.540$ |
| Depression, *n* (%) | 163 (17.91%) | 298 (6.07%) | $\chi^2 = 147.806, p < 0.001^{***}$ |
| Anxiety, *n* (%) | 92 (10.11%) | 162 (3.30%) | $\chi^2 = 85.416, p < 0.001^{***}$ |
| Psychotic-like experiences, *n* (%) | 355 (39.01%) | 710 (14.45%) | $\chi^2 = 309.885, p < 0.001^{***}$ |
| Low family functioning, *n* (%) | 697 (76.59%) | 1835 (37.35%) | $\chi^2 = 481.149, p < 0.001^{***}$ |
| Low psychological resilience, *n* (%) | 471 (51.76%) | 1065 (21.68%) | $\chi^2 = 357.747, p < 0.001^{***}$ |

Note: FHMD, family history of mental disorders; OP/OC, overprotection/overcontrol; SD, standard deviation; \*\*\*, $p < 0.001$ ; \*\*, $p < 0.01$ ; \*, $p < 0.05$ .

### 3.3. Results of binary logistic regression analysis

As shown in **Table 3** and **Figure 1A**, after controlling for confounding factors in the logistic regression model, the following factors remained independently associated with OP/OC experiences: being male (odds ratio 1.973, 95% confidence interval 1.685-2.311, corrected *p* < 0.001), being a single child (odds ratio 1.232, 95% confidence interval 1.033-1.471, corrected *p* = 0.046), having depression (odds ratio 1.436, 95% confidence interval 1.105-1.866, corrected *p* = 0.018), having psychotic-like experiences (odds ratio 2.231, 95% confidence interval 1.872-2.659, corrected *p* < 0.001), having low family functioning (odds ratio 3.808, 95% confidence interval 3.188-4.549, corrected *p* < 0.001), and having low psychological resilience (odds ratio 2.126, 95% confidence interval 1.799-2.511, corrected *p* < 0.001). There were no statistically significant associations between OP/OC and the following factors: province, age, years of education, ethnicity, parental separation, left-behind experiences, family history of mental disorders, and anxiety (all corrected *p* > 0.05), after controlling for confounding factors.

**Figure 1.**
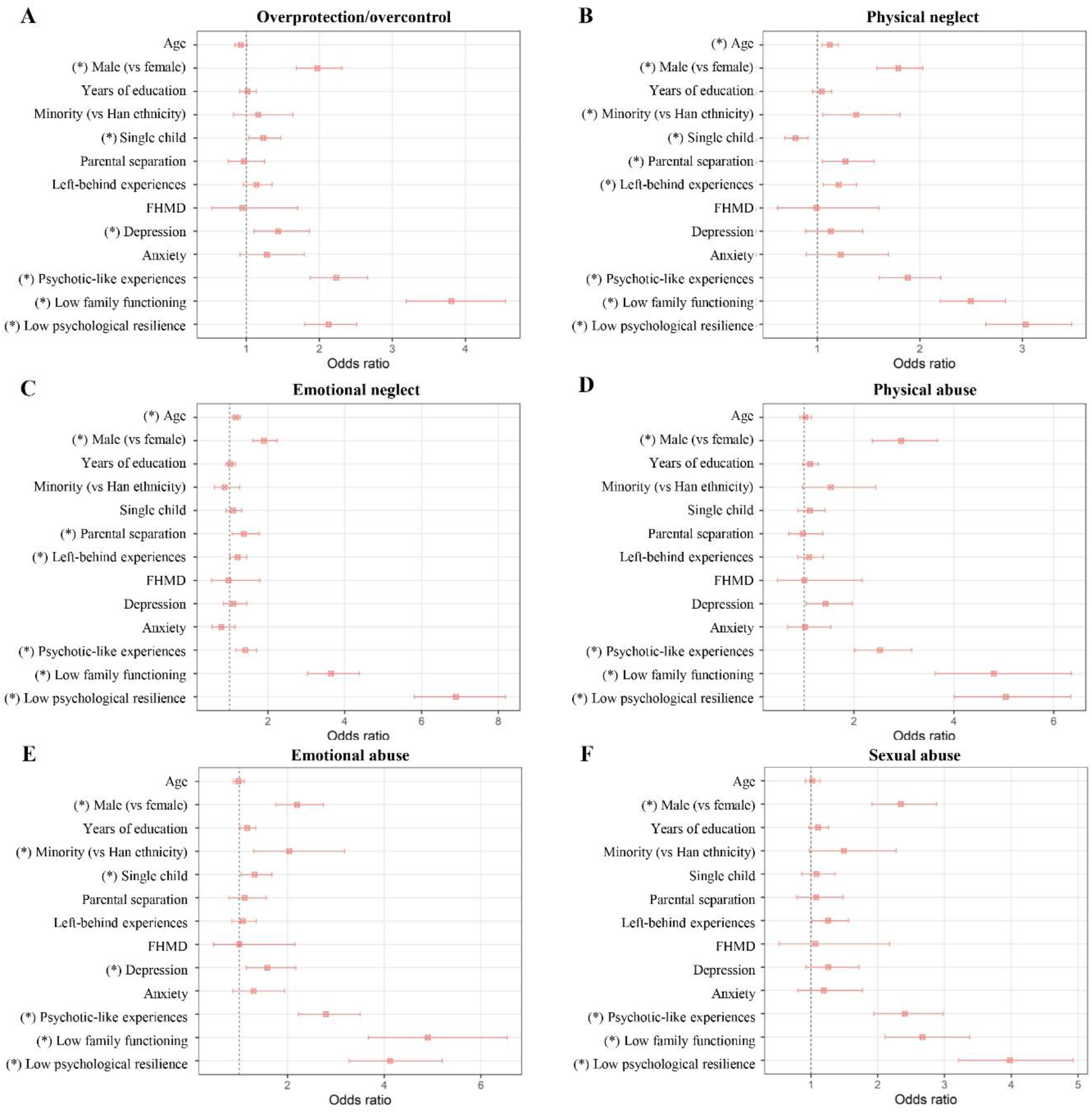
Results of separate binary logistic regression analyses for factors associated with OP/OC (**A**) and for factors associated with other trauma subtypes (**B-F**). The odds ratios with 95% confidence intervals are presented, and the “*” indicates a significant association with corrected *p* < 0.05. FHMD, family history of mental disorder; OP/OC, overprotection/overcontrol.

**Table 3.** Results of the binary logistic regression analysis for factors associated with OP/OC. The presented *p* values were FDR-corrected.

| Variables | B | SE | Wald | Significance | Odds ratio | 95% CI for odds ratio |  |
| --- | --- | --- | --- | --- | --- | --- | --- |
|  |  |  |  |  |  | Lower | Upper |
| Age | -0.080 | 0.046 | 3.055 | $p = 0.149$ | 0.923 | 0.844 | 1.010 |
| Male (vs female) | 0.680 | 0.081 | 70.915 | $p < 0.001^{***}$ | 1.973 | 1.685 | 2.311 |
| Years of education | 0.017 | 0.056 | 0.092 | $p = 0.849$ | 1.017 | 0.911 | 1.136 |
| Minority (vs Han ethnicity) | 0.150 | 0.175 | 0.738 | $p = 0.507$ | 1.162 | 0.825 | 1.637 |
| Single child | 0.209 | 0.090 | 5.368 | $p = 0.046^*$ | 1.232 | 1.033 | 1.471 |
| Parental separation | -0.033 | 0.130 | 0.065 | $p = 0.849$ | 0.968 | 0.750 | 1.248 |
| Left-behind experiences | 0.131 | 0.088 | 2.215 | $p = 0.218$ | 1.140 | 0.959 | 1.354 |
| FHMD | -0.057 | 0.301 | 0.036 | $p = 0.849$ | 0.944 | 0.524 | 1.702 |
| Depression | 0.362 | 0.134 | 7.323 | $p = 0.018^*$ | 1.436 | 1.105 | 1.866 |
| Anxiety | 0.247 | 0.172 | 2.067 | $p = 0.218$ | 1.281 | 0.914 | 1.795 |
| Psychotic-like experiences | 0.803 | 0.089 | 80.433 | $p < 0.001^{***}$ | 2.231 | 1.872 | 2.659 |
| Low family functioning | 1.337 | 0.091 | 217.369 | $p < 0.001^{***}$ | 3.808 | 3.188 | 4.549 |
| Low psychological resilience | 0.754 | 0.085 | 78.765 | $p < 0.001^{***}$ | 2.126 | 1.799 | 2.511 |

Note: CI, confidence interval; FDR, false discovery rate; FHMD, family history of mental disorder; OP/OC, overprotection/overcontrol; SE, standard error; \*\*\*, $p < 0.001$ ; \*\*, $p < 0.01$ ; \*, $p < 0.05$ .

### 3.3. Supplementary analyses on other trauma subtypes

As shown in **Table 4**, significant positive correlations were found between the OP/OC score and scores of all other trauma subtypes in the CTQ-33 (all *p* < 0.001), confirming that OP/OC is highly positively associated with the other trauma subtypes. Results of the separate binary logistic regression analyses for factors associated with other trauma subtypes are shown in **Figures 1B-1F**. Generally, it was found that the OP/OC and other trauma subtypes have both shared and unique associated factors (*p* < 0.05 after corrections). For example, all trauma subtypes including OP/OC were found to be positively associated with having psychotic-like experiences, having low family functioning, and having low psychological resilience; meanwhile, parental separation was found to be associated with only the physical neglect and emotional neglect experiences (**Figure 1**).

**Table 4.** Spearman correlation coefficients between the OP/OC score and scores of other trauma subtypes in the CTQ-33.

|  | Physical<br>neglect | Emotional<br>neglect | Physical<br>abuse | Emotional<br>abuse | Sexual<br>abuse |
| --- | --- | --- | --- | --- | --- |
| OP/OC | 0.340*** | 0.380*** | 0.408*** | 0.454*** | 0.353*** |
| Physical neglect |  | 0.551*** | 0.392*** | 0.404*** | 0.370*** |
| Emotional neglect |  |  | 0.343*** | 0.338*** | 0.285*** |
| Physical abuse |  |  |  | 0.559*** | 0.574*** |
| Emotional abuse |  |  |  |  | 0.450*** |

Note: CTQ-33, the 33-item Childhood Trauma Questionnaire; OP/OC, overprotection/overcontrol; the “\*” indicates a significant correlation with $p < 0.001$ .

### 3.4. Supplementary analyses on possible sex differences

Results of separate logistic regression analyses in the female or male participants are shown in **Figure 2**. Generally, most of the associated factors of OP/OC were found to be consistent across the female and male participants (corrected *p* < 0.05 in both the two subsamples). The exceptions were that OP/OC was found to be associated with depression in only the female participants, and associated with anxiety in only the male participants (**Figure 2**).

**Figure 2.**
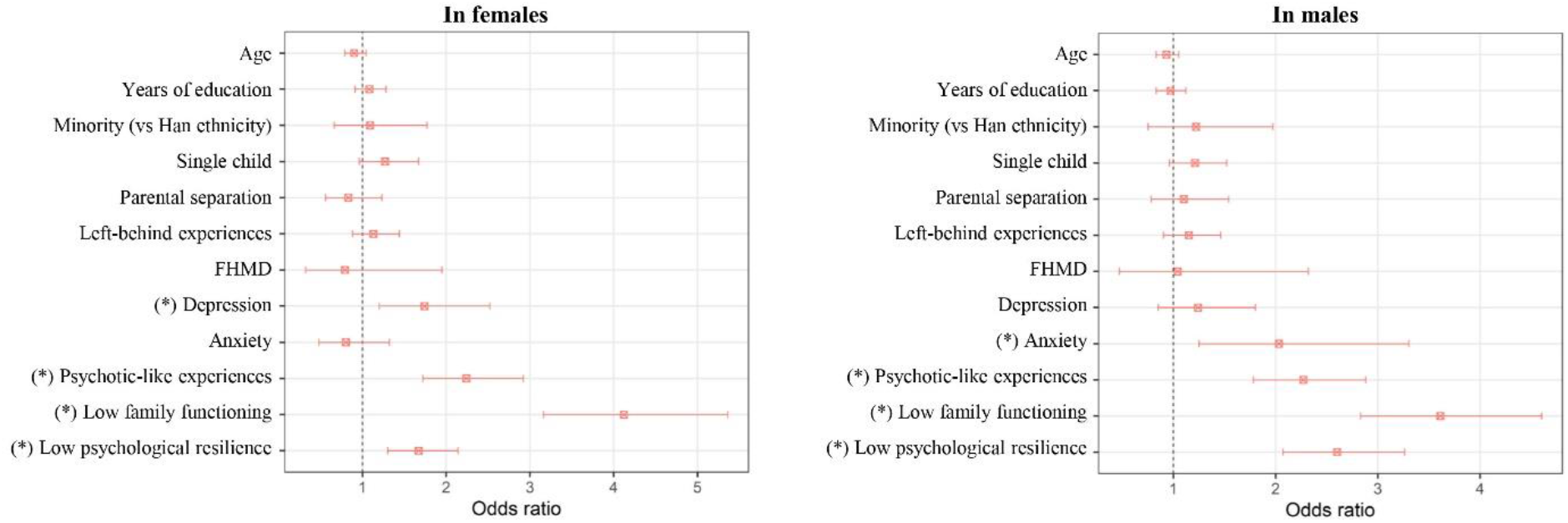
Results of separate binary logistic regression analyses for factors associated with OP/OC in the female or male participants. The odds ratios with 95% confidence intervals are presented, and the “*” indicates a significant association with corrected *p* < 0.05. FHMD, family history of mental disorder; OP/OC, overprotection/overcontrol.

### 3.5. Validation analysis

When using the cutoff points of OP/OC subscale score >= 12 or >= 14, 20.81% (1212/5823) and 12.31% (717/5823) of the surveyed participants were categorized as having OP/OC experiences, respectively. The following factors were still found to be significantly associated with OP/OC when using the above different cutoff points: being male, being a single child, having depression, having psychotic-like experiences, having lower family functioning, and having lower psychological resilience (all corrected *p* < 0.05).

## 4. Discussion

In this study, we investigated the possible associations between childhood OP/OC experiences and a series of socio-demographic and psychological factors in a nationwide sample of Chinese university students. Generally, our results indicated that being male, being a single child, having depression, having psychotic-like experiences, having lower family functioning, and having lower psychological resilience were independently associated with childhood OP/OC experiences. The OP/OC was also positively associated with all the other assessed trauma subtypes (abuses and neglects) in the CTQ-33. These results may provide initial evidence that childhood OP/OC experiences might have negative effects on the mental health in young populations.

In the current study, we firstly explored an appropriate cutoff point for the OP/OC subscale in CTQ-33 based on the statistical distributions in the surveyed sample. The cutoff was estimated at ≥ 13, and 15.63% (910/5823) of the surveyed participants were categorized as having OP/OC experiences according to such cutoff point. This prevalence is higher than those of physical abuse (8.59%, 500/5823), emotional abuse (8.07%, 470/5823) and sexual abuse (8.98%, 522/5823), but lower than those of physical neglect (33.52%, 1952/5823) and emotional neglect (16.26%, 947/5823) in the current sample. Note that all the identified associated factors of OP/OC were found to be unchanged when using different cutoff points at >= 12 and >= 14; therefore, the main conclusions in this study are unlikely to be largely driven by different choices in cutoff points.

Using the binary logistic regression model, we found that being male and being a single child are positively associated with childhood OP/OC experiences (**Table 3**). The observed sex effects on OP/OC are partly consistent with previous research showing sex differences in perceived parenting styles (Eun et al., 2018). We propose that several biological and social factors might account for such sex differences. For example, boys are favored over girls in traditional Chinese culture, which may lead to more focus on the boys than girls in some families. For the same reason, the children which are the single child of their family might attract more attention, and even overprotective parental strategies. Notably, we did not find significant associations between parental separation and OP/OC. One possible reason might be that children can be affected differently by whether their parents’ separation was amicable or conflict-ridden (Beal et al., 2019).

The regression analyses suggested that having depression is independently associated with OP/OC, even after adjusting for possible confounding effects of all other variables (**Table 3**). To our knowledge, the findings in previously published studies are not totally consistent regarding the possible associations between OP/OC experiences and levels of depressive symptoms in later life. For example, one earlier research reported a strong association between negative parenting behaviors such as overprotection and later depressive symptom (Williamson et al., 2017). However, there is also research suggesting that paternal overcontrols can predict lower depressive symptoms (Basili et al., 2021). It is noteworthy that compared to most of these studies, our study has a much larger sample size and thus a higher statistical power. Therefore, this study may provide more solid evidence in support of the positive association between OP/OC and depression.

Our results also suggested that having psychotic-like experiences is independently associated with OP/OC (**Table 3**). To the best of our knowledge, this study is one of the first reports to suggest a positive relationship between OP/OC and psychotic-like experiences. Psychotic-like experiences are subclinical delusion-like or hallucination-like symptoms, which are related to increased risks of developing subsequent mental disorders (Wu, Liu, Zhang, et al., 2022). Previous studies have shown that some other subtypes of childhood trauma such as abuses and neglects would strongly increase the risks of developing schizophrenia and other psychotic disorders (Chaiyachati & Gur, 2021; Vieira et al., 2020), which may be presented as having psychotic-like experiences in the early stage (Read et al., 2005). Here, our results suggest that OP/OC, as another subtype of childhood trauma, is also associated with psychotic-like experiences.

Additionally, we found that childhood OP/OC experiences are associated with lower family functioning and lower psychological resilience (**Table 3**). Both family dysfunction (Wiegand-Grefe et al., 2019) and decreased psychological resilience (Ungar & Theron, 2020) have been linked to higher risks of developing mental problems. Lower family functioning was also associated with lower well-being and higher risks of substance use (Mersky et al., 2013; Sitnick et al., 2014). These findings, together with the observed significant effects of OP/OC on depression and psychotic-like experiences, may thus highlight the unignorable negative effects of OP/OC experiences on young people’s mental health.

As supplementary analyses, we have explored the possible differences in associated factors between OP/OC and other childhood trauma subtypes. It was found that some associated factors, such as having psychotic-like experiences, lower family functioning, and lower psychological resilience, were shared for all different trauma subtypes including OP/OC (**Figure 1**). Some differences were also found: for example, being a single child was positively associated with OP/OC but negatively associated with physical neglect; furthermore, having depression was positively associated with OP/OC and emotional abuse, but not significantly associated with other trauma subtypes (**Figure 1**). Therefore, while being a subtype of traumatic experiences, there might be both common and unique features between the OP/OC and other trauma subtypes.

We have also explored the possible sex differences in relationships between OP/OC and other factors by performing analyses in the female and male participants separately. Generally, we found that most of the associated factors of OP/OC were consistent across the female and male participants; however, interestingly, the OP/OC experiences were associated with depression in only the female participants, and associated with anxiety in only the male participants (**Figure 2**). There has been ample evidence for significant sex differences in multiple psychological characteristics, e.g., that females are more likely to be affected by depression (Bangasser & Cuarenta, 2021; Rubinow & Schmidt, 2019). Here, our results may partly help to further understand the sex differences in these psychological characteristics in the aspect of different influences of childhood OP/OC experiences.

This study has certain limitations. First, because of the nature of cross-sectional survey, we are unable to establish the causality in relationships between OP/OC experiences and other factors. Therefore, further longitudinal studies are needed to address such a limitation. Second, several self-reported retrospective scales were used in this study, which may lead to memory-related biases. Third, the OP/OC experiences from one’s father, mother, or other family members were not distinguished in the CTQ-33, which might actually have different associated socio-demographic factors and psychological effects. This limitation may be overcome by using other scales in future studies. Last, while only healthy participants were included in the current study, further studies conducted in clinical populations with mental disorders may provide more important implications for understanding the negative effects of childhood OP/OC experiences.

## 5. Conclusions

In conclusion, this study investigated the possible associated factors of childhood OP/OC experiences in young populations using the CTQ-33 and a relatively large, nationwide sample of Chinese university students. The main findings include that being male, being a single child, having depression, having psychotic-like experiences, having lower family functioning, and having lower psychological resilience were independently associated with childhood OP/OC experiences. The OP/OC was also positively associated with all the other trauma subtypes (abuses and neglects) in the CTQ-33; nevertheless, the OP/OC and other subtypes of trauma were found to have both shared and unique associated factors. These results may provide initial evidence that childhood OP/OC experiences would have negative effects on young people’s mental health, and highlight the great values of further investigations on OP/OC especially in clinical populations.

## Data Availability

All data produced in the present study are available upon reasonable request to the authors.

## Author contributions

JZ, ZW, ZL and YL contributed to the conception and design of the study. JZ, ZW, MC, MY, LZ, MS, DL, GC, QY, HT, CA, ZL and YL contributed to the data acquisition. JZ, ZW and YL contributed to the analysis and interpretation of data. JZ and YL drafted the manuscript. HT, MS and XH revised the manuscript. All authors read and approved the final manuscript.

## Acknowledgments

We would like to thank all students who served as research participants.

